# Eye care practitioners and falls prevention: protocol for a scoping review

**DOI:** 10.1101/2024.04.22.24306149

**Authors:** Jingyi Chen, Khyber Alam, Si Ye Lee, Anne-Marie Hill

**Affiliations:** Department of Optometry, Health and Medical Sciences, The University of Western Australia, Perth, Western Australia, Australia; WA Centre for Health & Ageing, The University of Western Australia Perth, Western Australia, Australia; School of Allied Health, The University of Western Australia, Perth, Western Australia, Australia

**Keywords:** accidental falls, falls prevention, older adults, vision, optometry

## Abstract

**Introduction:** Falls are the leading cause of injury in older adults and can lead to substantial costs for the individual and society. Eye care practitioners in the community provide services for a large proportion of the older adult population and can play a key role in falls prevention, however it is unclear whether they are implementing evidence-based recommendations in practice. The aim of this scoping review is to map and synthesise the current evidence for community-based eye care practitioners translating falls prevention evidence into clinical practice.

**Methods and analysis:** The study will use the framework by Arksey and O’Malley and the Preferred Reporting Items for Systematic Reviews and Meta-Analyses Extension for Scoping Reviews (PRISMA-ScR). Studies that have been published in English from 1990 to current on the topic of eye care practitioners and falls prevention in older adults will be searched on MEDLINE, Embase, and CINAHLPlus databases. Grey literature will be searched using Web of Science and OpenMD. The titles and abstracts of results will be screened against the inclusion criteria and full texts will be analysed. Data from final included articles will be extracted by two independent reviewers using a tool created according to the Joanna Briggs Institute guidelines. The World Falls Guidelines will be used as a framework for data mapping. Results will be charted, collated, and summarised narratively.

**Ethics and dissemination:** It is anticipated that the results from this scoping review will highlight any gaps in the literature regarding eye care practitioner awareness and implementation of falls prevention guidelines and inform future research and clinical recommendations. Ethics approval is not required. Findings will be submitted to a peer-reviewed journal for publication.

## Introduction

Falls are common in older people and are the leading cause of injury for adults 65 years and over^1, 2^. Falls can have significant costs to the individual, their caregivers, and the healthcare system.^3, 4^ In the United States (US), approximately $50 billion is spent on medical costs related to falls in older adults.^5^ Advancing age is a risk factor^6^ and the incidence of falls worldwide is expected to continue to increase due to the ageing population^7^.

In the US, around 60% of people with vision impairment are aged 65 or over.^8^ Given that many older adults report having an eye examination^9^, community eye care practitioners can play a key role in falls prevention as part of a multidisciplinary care approach. Each interaction with an older adult provides an opportunity whereby eye care practitioners can offer intervention, education, or referral to patients in accordance with evidence-based falls recommendations. Whilst there is evidence which suggests that some eye care interventions, such as cataract surgery, will reduce falls risk^10^, other evidence suggests that certain interventions may increase falls risk^11, 12^. This highlights the importance of eye care practitioners understanding falls prevention evidence to ensure they can make informed recommendations to patients.

The World Falls Guidelines (WFG)^13^ were developed for use by healthcare and other professionals working with older adults, which encompasses eye care practitioners. It incorporates recent evidence of falls prevention into a framework with a patient-centred approach.^13^ The guidelines include broad recommendations that are relevant to all healthcare practitioners as well as specific recommendations for vision. Falls prevention management by community eye care practitioners has the potential to reduce the growing number of older adult falls and consequent injuries, however there is scant evidence for whether this is occurring in practice.

A preliminary search of MEDLINE and the Cochrane Database of Systematic Reviews was conducted which found no current systematic or scoping reviews on the topic of falls prevention implementation and community eye care practitioners. As scoping studies incorporate a range of study designs, they are helpful for topics with emerging evidence as the paucity of randomised controlled trials makes it difficult to undertake systematic reviews.^14^ Scoping reviews can assist in the mapping of available evidence to identify gaps in the literature.^14^ The proposed scoping review aims to bring together a broad range of sources to map the evidence for eye care practitioners’ awareness and implementation of falls prevention guidelines, and identify any gaps in the literature.

## Research question

The research question will be “What is the evidence in current literature regarding whether eye care practitioners are aware of and implementing evidence-based practice for falls prevention in the community?”

## Eligibility criteria

In accordance with Joanna Briggs Institute, the inclusion and exclusion criteria will be defined by ‘participants’, ‘concept’ and ‘context’.^15^ The participants will be eye care practitioners, which for this review will include optometrists, who deliver primary care, and ophthalmologists, who deliver secondary and tertiary eye care. The concept is eye care practitioners’ knowledge, awareness, and behaviours of implementing evidence-based falls recommendations. The context for this review will be community eye care practices, therefore randomised-control trials and studies conducted in hospital and residential aged-care facility settings will not be included. Studies will be excluded if they were published before 1990 to ensure recency, or if they were not published in English. This scoping review will consider all published study types, as well as grey literature, to capture a wide range of information.

## Methods and Analysis

This review will be conducted in accordance with the Preferred Reporting Items for Systematic Reviews and Meta-Analyses Extension for Scoping Reviews (PRISMA-ScR)^16^ in conjunction with the Arksey and O’Malley framework for scoping reviews^17^. The WFG^13^ will be used as a framework to map and chart the available evidence.

### Search strategy and evidence selection

An initial search of MEDLINE was undertaken to identify relevant articles. Keywords and index terms were used to develop a full search strategy with the guidance of an experienced librarian. An example search can be found in Supplementary File 1. The databases to be searched include MEDLINE, Embase, and CINAHLPlus, with the search strategy adapted for each database. The organisations listed on the Ophthalmology directory on OpenMD and Web of Science will be searched for grey literature.

All identified citations from the search will be uploaded to EndNote 21 for screening. Duplicates will be removed, then titles and abstracts will be screened against the inclusion criteria by two independent reviewers. The full text articles will be retrieved and assessed against the inclusion criteria. Any disagreements between reviewers will be resolved through discussion, and a third reviewer if needed. The search strategy results will be presented in a PRISMA-ScR flow diagram^16^.

### Data Extraction

A data extraction tool will be developed by the reviewers and include details about author, publication year, country, study design, participant characteristics, and a summary for each study. Through the data extraction process, additional variables may be added to the tool if necessary and documented. Any quotes that pertain to or describe eye care practitioners’ falls prevention evidence awareness or behaviour will be extracted.

### Data Collation and Presentation

Data mapping will be completed by two reviewers with a third reviewer as required to seek consensus. Using the WFG as a framework, data will be charted with the sub-sections of i) falls risk stratification, ii) assessment, and iii) management and intervention. Results will be presented in narrative summary as well as tables and diagrams where appropriate.

## Data Availability

All data produced are available online

## Declarations

The authors declare there are no conflicts of interest.

## Contributors

JC, KA, and A-MH contributed to the study design and formulation of the research objectives. JC and AM-H wrote the protocol that was reviewed by all authors. All authors edited and approved the protocol prior to submission.

## Funding

Anne-Marie Hill is supported by a National Health and Medical Research Council (NHMRC) of Australia Investigator (EL2) award (GNT1174179) and the Royal Perth Hospital Research Foundation.

Si Ye Lee is supported by a Perth Eye Foundation – NHMRC HDR Scholarship award through the University of Western Australia (UWA) and the Australian Government Research Training Program (RTP) Fees Offset scholarship.

## Notes

### Competing Interest Statement

The authors have declared no competing interest.

## References

1. Moncada LVV, Mire LG. Preventing Falls in Older Persons. Am Fam Physician. 2017;96(4):240–7.

2. Bergen G, Stevens MR, Burns ER. Falls and fall injuries among adults aged≥ 65 years— United States, 2014. Morbidity and Mortality Weekly Report. 2016;65(37):993–8.

3. Heinrich S, Rapp K, Rissmann U, Becker C, König HH. Cost of falls in old age: a systematic review. Osteoporosis International. 2010;21(6):891–902.

4. Davis JC, Robertson MC, Ashe MC, Liu-Ambrose T, Khan KM, Marra CA. International comparison of cost of falls in older adults living in the community: a systematic review. Osteoporosis International. 2010;21(8):1295–306.

5. Centers for Disease Control and Prevention. Older Adult Falls Data 2023 [cited 2024 Mar 8]. Available from: https://www.cdc.gov/falls/data/index.html.

6. Ambrose AF, Paul G, Hausdorff JM. Risk factors for falls among older adults: A review of the literature. Maturitas. 2013;75(1):51–61.

7. Tricco AC, Thomas SM, Veroniki AA, Hamid JS, Cogo E, Strifler L, et al. Comparisons of Interventions for Preventing Falls in Older Adults: A Systematic Review and Meta-analysis. JAMA. 2017;318(17):1687–99.

8. Killeen OJ, De Lott LB, Zhou Y, Hu M, Rein D, Reed N, et al. Population Prevalence of Vision Impairment in US Adults 71 Years and Older: The National Health and Aging Trends Study. JAMA Ophthalmol. 2023;141(2):197–204.

9. Ehrlich JR, Ndukwe T, Solway E, Woodward MA, Singer DC, Newman-Casey PA, et al. Self-reported Eye Care Use Among US Adults Aged 50 to 80 Years. JAMA Ophthalmol. 2019;137(9):1061–1066.

10. Keay L, Ho KC, Rogers K, McCluskey P, White AJR, Morlet N, et al. The incidence of falls after first and second eye cataract surgery: a longitudinal cohort study. Medical Journal of Australia. 2022;217(2):94–9.

11. Cumming RG, Ivers R, Clemson L, Cullen J, Hayes MF, Tanzer M, Mitchell P. Improving vision to prevent falls in frail older people: a randomized trial. J Am Geriatr Soc. 2007;55(2):175–81.

12. Meuleners LB, Lee AH, Ng JQ, Morlet N, Fraser ML. First eye cataract surgery and hospitalization from injuries due to a fall: a population-based study. J Am Geriatr Soc. 2012;60(9):1730–3.

13. Montero-Odasso M, van der Velde N, Martin FC, Petrovic M, Tan MP, Ryg J, et al. World guidelines for falls prevention and management for older adults: a global initiative. Age and Ageing. 2022;51(9).

14. Peters MDJ, Godfrey C, McInerney P, Munn Z, Tricco AC, Khalil H. Scoping reviews. Joanna Briggs Institute reviewer’s manual. 2017;2015:1–24.

15. Peters MDJ, Godfrey CM, Khalil H, McInerney P, Parker D, Soares CB. Guidance for conducting systematic scoping reviews. JBI Evidence Implementation. 2015;13(3):141–6.

16. Tricco AC, Lillie E, Zarin W, O’Brien KK, Colquhoun H, Levac D, et al. PRISMA Extension for Scoping Reviews (PRISMA-ScR): Checklist and Explanation. Ann Intern Med. 2018;169(7):467–73.

17. Arksey H, O’Malley L. Scoping studies: towards a methodological framework. Int J Soc Res Methodol. 2005;8(1):19–32.

