## Supplementary File 1 for "Eye care practitioners and falls prevention: protocol for a scoping review"

MEDLINE Literature Example Search Strategy

Database: Ovid MEDLINE(R), Ovid MEDLINE(R) In-Process & Other Non-Indexed Citations, Ovid MEDLINE(R) Daily and Ovid OLDMEDLINE(R) <1990 to Present>

Search Strategy:

1. (optometr* or optic* or ophthalmolog* or eyecare professional or eyecare practitioner or eye care profession* or eye care practitioner).tw.
2. Optometrists/ or optometry/ or ophthalmology/ or Ophthalmologists/
3. 1 or 2
4. fall*.tw.
5. Accidental Falls/
6. 4 or 5
7. exp aged/ or exp geriatrics/ or exp geriatric nursing/ or (centarian* or centenarian* or elder* or eldest or frail* or geriatri* or nonagenarian* or octagenarian* or octogenarian* or old age* or older adult* or older age* or older female* or older male* or older man or older men or older patient* or older people or older person* or older population or older subject* or older woman or older women or oldest old* or senior* or senium or septuagenarian* or supercentenarian* or very old*).tw.
8. Aged/ or "Aged, 80 and over"/
9. 7 or 8
10. 3 and 6 and 9
